# Long-Term Clinical Outcomes of Sotatercept in Pulmonary Hypertension: A Retrospective Cohort Analysis

**DOI:** 10.1101/2025.08.05.25333015

**Authors:** Azka Naeem, Olayiwola Bolaji, George G. Kidess, Joud Fahed, Sultana Jahan, Muhammad Hashim Khan, Shaunak Mangeshkar, Steven Sorci, Hayah Kassis-George, Sourbha Dani, Chadi Alraies

## Abstract

**Background:** Sotatercept, an activin receptor ligand trap, has demonstrated significant short-term hemodynamic improvements in pulmonary hypertension (PH) based on clinical trials. However, evidence regarding long-term outcomes in real-world settings remains limited.

**Methods:** Using data from the TriNetX global federated health research network covering 102 healthcare organizations, we conducted a retrospective cohort study comparing 185 patients with PH receiving sotatercept to 212,349 PH patients not receiving sotatercept. After 1:1 propensity score matching to balance baseline characteristics, we assessed outcomes at 6 months, 1 year, 3 years, and 5 years post-initiation and cox proportional hazards was used. Primary outcomes included all-cause mortality, rehospitalization rates, major adverse cardiovascular events (MACE), and BNP elevation (≥100 pg/mL).

**Results:** Over 5 years of follow-up, sotatercept treatment was associated with significantly lower all-cause mortality (5.4% vs. 27.3%; risk difference, -21.9 percentage points [95% CI, -29.1 to -14.7]; p<0.001; hazard ratio [HR], 0.40 [95% CI, 0.16-0.99]). Rehospitalization rates were also substantially reduced in the sotatercept group (15.4% vs. 38.1%; risk difference, -22.8 percentage points [95% CI, - 35.8 to -9.7]; p=0.002; HR, 0.12 [95% CI, 0.03-0.49]). MACE occurrence was significantly lower with sotatercept (6.4% vs. 15.5%; risk difference, -9.1 percentage points [95% CI, -16.2 to -2.0]; p=0.011; HR, 0.45 [95% CI, 0.14-1.41]), while BNP elevation showed no significant difference (12.8% vs. 10.6%; p=0.626). Treatment benefits were observed as early as 6 months post-initiation, progressively increasing magnitude through 5 years of follow-up. Sensitivity analyses using alternative matching methods and instrumental variable approaches confirmed the robustness of these findings.

**Conclusions:** In this large real-world cohort study with long-term follow-up, sotatercept was associated with substantial reductions in mortality, rehospitalization, and cardiovascular events in patients with pulmonary hypertension. These benefits increased over time, suggesting potential disease-modifying effects beyond acute hemodynamic improvements. These findings complement data from randomized controlled trials and support the role of sotatercept in improving long-term outcomes in pulmonary hypertension.

## Introduction

Pulmonary hypertension (PH) affects approximately 1% of the global population and up to 10% of individuals over 65 years of age with higher rates in females and older adults[1]. PH is a major contributor to morbidity and mortality. It is considered a complex medical diagnosis due to several interrelated factors. The clinical presentation is often nonspecific leading to frequent delays and misdiagnosis. The diagnostic process requires a stepwise approach: initial suspicion is typically raised by echocardiography with definitive diagnosis requiring right heart catheterization, which is essential to distinguish between pre-capillary and post-capillary forms and to guide management [2].

PH is consistently associated with increased mortality, with an age-standardized mortality rate for PAH of 0.27 per 100,000 globally [3]. It is an independent risk factor for adverse outcomes, particularly in the context of left-sided heart disease, chronic lung disease, and perioperative settings. The American Heart Association emphasizes the need for careful classification and risk assessment, especially in perioperative settings, due to the high risk of morbidity and mortality. PH is also poor prognostic marker, and its presence significantly worsens outcomes, including right heart failure, which is the leading cause of death in these patients. The economic burden of PH is substantial, driven by frequent hospitalizations, advanced therapies, and the need for specialized care.

Given the morbidity and mortality associated with pulmonary hypertension with economic implications,, novel target therapies have been developed. Medications for pulmonary arterial hypertension (PAH) target three main pathways: the endothelin pathway (endothelin receptor antagonists such as ambrisentan, bosentan, macitentan), the nitric oxide pathway (phosphodiesterase-5 inhibitors such as sildenafil and tadalafil, and soluble guanylate cyclase stimulators such as riociguat), and the prostacyclin pathway (prostacyclin analogues such as epoprostenol, treprostinil, iloprost). These agents, alone or in combination, improve exercise capacity, hemodynamics, and delay clinical worsening, but long-term survival remains suboptimal, with 5-year survival rates around 60% despite maximal therapy.[4] Sotatercept is a novel, first-in-class activin signaling inhibitor that targets pulmonary vascular remodeling, a mechanism distinct from vasodilation. Sotatercept, one of the novel drugs, demonstrated significant short-term hemodynamic improvements through modulation of the transforming growth factor beta (TGF-β) pathway . The need for sotatercept arose from persistent morbidity and mortality in PAH despite maximal use of established therapies, as highlighted by the lack of substantial survival improvement over the past decade. Phase 2 and 3 trials (PULSAR, STELLAR, and ZENITH) have demonstrated that sotatercept, when added to background therapy, significantly improves exercise capacity, pulmonary vascular resistance, NT-proBNP levels, and reduces the risk of clinical worsening and major events (including death, lung transplantation, and hospitalization) even in patients with advanced disease on maximal therapy [5,6]. Hence, in patients resistant to standard therapy, sotatercept was used as a third agent in WHO functional class II-III and a fourth in WHO class IV PAH [7,8].

However, evidence regarding long-term outcomes in real-world settings remain limited. Given the robust efficacy data and the unmet need in PH, increased use of sotatercept is expected in the future. This anticipated uptake, combined with the need to understand long-term safety, durability of benefit, and cost-effectiveness, highlights why there is a critical need to explore long-term outcomes in diverse patient populations. Hence, we conducted a retrospective cohort study using data from TriNetX’s global federated health network to assess primary outcomes, including all-cause mortality, rehospitalization rates, major adverse cardiovascular events (MACE), and BNP elevation at 6-month, 1-year, 3 years, and 5-year post-initiation.

## Methods

### Data Source and Study Population

We conducted a retrospective cohort analysis using data from the TriNetX global federated health research network, which provides access to electronic medical records from 102 healthcare organizations (HCOs). The network covers approximately 88 million patients across the United States, Europe, and Asia, allowing for analysis of real-world treatment patterns and outcomes [9,10]. This study was executed by the Declaration of Helsinki, and data were de-identified in compliance with the Health Insurance Portability and Accountability Act (HIPAA). The TriNetX platform has been validated for observational research in multiple therapeutic areas and has demonstrated high concordance with findings from randomized controlled trials [11,12].

### Study Design and Cohort Definition

Patients with pulmonary hypertension were identified using the International Classification of Diseases, Tenth Revision, Clinical Modification (ICD-10-CM) codes I27.0 (primary pulmonary hypertension) and I27.21 (secondary pulmonary arterial hypertension). We included adult patients (≥18 years) with diagnostic codes for pulmonary hypertension between January 1, 2019, and December 31, 2024. The index date for each patient was defined as the first recorded diagnosis of pulmonary hypertension or the first prescription of sotatercept, whichever occurred later.

Two cohorts were established: (1) patients with pulmonary hypertension who received sotatercept (RxNorm code 2678930) and (2) patients with pulmonary hypertension who did not receive sotatercept. Patients were required to have at least one healthcare encounter in the 12 months preceding the index date to ensure adequate baseline data for propensity score calculation. We excluded patients who had received other investigational pulmonary hypertension therapies or had less than 30 days of follow-up after the index date.

### Propensity Score Matching

We used propensity score matching to create balanced comparison groups to address potential selection bias and control for confounding factors [13]. Propensity scores were calculated using multivariable logistic regression that incorporated the following covariates: age, sex, race, ethnicity, comorbidities (heart failure, chronic obstructive pulmonary disease, diabetes mellitus, essential hypertension, tobacco use), concomitant medications (sildenafil, bosentan), and baseline left ventricular ejection fraction.

We performed 1:1 matching without replacement using a greedy nearest-neighbor algorithm with a caliper width of 0.2 standard deviations of the logit of the propensity score, as recommended by Austin et al. [14]. The matching quality was assessed by calculating standardized differences for each covariate before and after matching, with standardized differences <0.1 indicative of good balance between cohorts [15]. Density plots of propensity score distributions were generated to confirm the improved overlap after matching visually.

### Outcome Definitions and Follow-up

The primary outcomes of interest were all-cause mortality, rehospitalization rates, major adverse cardiovascular events (MACE), and elevated B-type natriuretic peptide (BNP) levels. All-cause mortality was determined using death records within the TriNetX system. Rehospitalization was defined using Current Procedural Terminology (CPT) codes for subsequent hospital inpatient or observation care (CPT 99231, 99232, 99233, 99462, and 1013668). MACE was a composite endpoint comprising cardiac arrest (ICD-10-CM I46, I46.9), cerebral infarction (I63, I63.50), and myocardial infarction (I21, I21.3, I21.4, I21.9, I21.A, I21.A1). As recorded in laboratory data, BNP elevation was defined as values ≥100 pg/mL.

The time window for outcome assessment began one day after the index event. It extended through the duration of available follow-up data, with analyses conducted at predefined time points of six months, one year, three years, and five years. Patients were censored at the time of their last recorded healthcare encounter in the database if they did not experience the outcome of interest during the follow-up period.

### Statistical Analysis

For each outcome, we performed both risk analysis and survival analysis. Risk analysis calculated the proportion of patients experiencing each outcome within the specified time windows. We reported risk differences with 95% confidence intervals (CI) and corresponding p-values. We used the Kaplan-Meier method for survival analysis to estimate event-free survival probabilities and median time to event. Log-rank tests were performed to compare the survival distributions between the two cohorts. Hazard ratios (HR) with 95% CI were estimated using Cox proportional hazard models. The proportional hazards assumption was verified using Schoenfeld residuals and formal testing [16].

We also analyzed instances of rehospitalization, MACE, and BNP elevation outcomes to assess the frequency of events per patient. Patients with prior occurrences of the outcomes before the start of the time window were excluded from the respective analyses to ensure that only new-onset events were captured. We report the mean, standard deviation, and median number of events per patient for each analysis, along with appropriate statistical tests for between-group comparisons.

All statistical analyses were conducted using the TriNetX Analytics features, which implement established methods for propensity score matching and time-to-event analyses [17]. A two-sided p-value <0.05 was considered statistically significant. To address the issue of multiple comparisons across different outcomes and time points, we applied the Benjamini-Hochberg procedure to control the false discovery rate at 0.05 [18].

### Sensitivity Analyses

We performed several sensitivity analyses to assess the robustness of our findings. First, we conducted an analysis using different matching algorithms (optimal matching and matching with replacement) to determine if the choice of matching method affected the results. Second, we implemented a high-dimensional propensity score method incorporating additional variables extracted from diagnostic codes, procedures, and medications to address potential unmeasured confounding [19]. Third, we performed an analysis restricted to patients with primary pulmonary hypertension (ICD-10-CM I27.0) to evaluate treatment effects in a more homogeneous population. Finally, we conducted an instrumental variable analysis using healthcare facility preference for sotatercept as an instrument to address potential unmeasured confounding [20] further.

## Results

### Study Population and Baseline Characteristics

After propensity score matching, 185 patients with pulmonary hypertension receiving sotatercept were compared with 185 matched controls not receiving sotatercept. Baseline characteristics were well-balanced between groups after matching (standardized differences <0.15 for all variables), including demographics, comorbidities, and concomitant medications (Table 1). The mean age was 53.1 ± 15.4 years in the sotatercept group and 52.1 ± 20.3 years in the control group (p=0.592). Both groups had a predominance of female patients (77.8% vs. 76.2%, p=0.711) and similar racial distribution (73.0% White in the sotatercept group vs 75.7% in the control group, p=0.552). Heart failure was the most common comorbidity in both groups (80.0% vs 78.9%, p=0.797).

**Table 1.** Baseline Characteristics Before and After Propensity Score Matching.

| Baseline Characteristics of the Study Cohort Before and After Propensity Score Matching Based on Sotatercept_in_pHTN |  |  |  |  |  |  |
| --- | --- | --- | --- | --- | --- | --- |
|  | Before Propensity Matching |  |  | After Propensity Matching |  |  |
|  | Pulmonary HTN on sotatercept (n = 185) | Pulmonary HTN not on Sotatercept (n= 212, 347) | SMD | Pulmonary HTN on sotatercept (n = 185) | Pulmonary HTN not on Sotatercept (n = 185) | SMD |
| **Demographics** |  |  |  |  |  |  |
| Age (Mean ± SD), yrs | 53.1 +/- 15.4 | 62.5 +/- 20.6 | 0.521 | 53.1 +/- 15.4 | 52.1 +/- 20.3 | 0.056 |
| Female | 144 (77.8%) | 115,168 (58.1%) | 0.432 | 144 (77.8%) | 141 (76.2%) | 0.039 |
| Male | 41 (22.2%) | 74,899 (37.8%) | 0.346 | 41 (22.2%) | 44 (23.8%) | 0.039 |
| White | 135 (73.0%) | 118,832 (60.0%) | 0.278 | 135 (73.0%) | 140 (75.7%) | 0.062 |
| Black | 18 (9.7%) | 39,581 (20.0%) | 0.291 | 18 (9.7%) | 18 (9.7%) | <0.001 |
| Hispanic or Latino | 22 (11.9%) | 12,631 (6.4%) | 0.192 | 22 (11.9%) | 14 (7.6%) | 0.146 |
| Non-Hispanic/Latino | 155 (83.8%) | 140,988 (71.2%) | 0.306 | 155 (83.8%) | 164 (88.6%) | 0.141 |
| **Comorbidities** |  |  |  |  |  |  |
| Hypertension | 108 (58.4%) | 101,875 (51.4%) | 0.140 | 108 (58.4%) | 105 (56.8%) | 0.033 |
| Diabetes mellitus | 39 (21.1%) | 53,275 (26.9%) | 0.136 | 39 (21.1%) | 29 (15.7%) | 0.140 |
| Chronic obstructive pulmonary disease | 47 (25.4%) | 43,328 (21.9%) | 0.083 | 47 (25.4%) | 48 (25.9%) | 0.012 |
| Tobacco use | 12 (6.5%) | 8,109 (4.1%) | 0.107 | 12 (6.5%) | 12 (6.5%) | <0.001 |
| Heart failure | 148 (80%) | 73,641( 37.2%) | 0.966 | 148 (80%) | 146 (78.9%) | 0.027 |
| **Medication** |  |  |  |  |  |  |
| Endothelin receptor antagonists | 12 (6.5%) | 1,412 (0.7%) | 0.314 | 12 (6.5%) | 10 (5.4%) | 0.046 |
| Medications increasing cGMP | 114 (61.6%) | 15,168 (7.7%) | 1.377 | 114 (61.6%) | 116 (62.7%) | 0.022 |
| Others |  |  |  |  |  |  |
| Left Ventricular Ejection Fraction (LVEF) (%) | 33 (17.8%) | 24,596 (12.4%) | 0.507 | 33 (17.8%) | 33 (17.8%) | 0.446 |
| 40 - 50 % | 10 (5.4%) | 4,070 (2.1%) | 0.178 | 10 (5.4%) | 10 (5.4%) | <0.001 |
| 50 - 0 % | 32 (17.3%) | 21,473 (10.8%) | 0.187 | 32 (17.3%) | 28 (15.1%) | 0.059 |

Concomitant pulmonary hypertension therapies were balanced between groups, with 61.6% of sotatercept patients and 62.7% of controls receiving sildenafil (p=0.830) and 6.5% and 5.4% receiving bosentan (p=0.660), respectively.

### Follow-up and Clinical Outcomes

Median follow-up was 145 days (interquartile range [IQR], 146 days) in the sotatercept group and 894 days (IQR, 1583 days) in the control group after matching. The differential follow-up duration necessitated time-to-event analyses to account for varying exposure periods.

### All-Cause Mortality

All-cause mortality was significantly lower in the sotatercept group compared with the control group at 5 years of follow-up (5.4% vs. 27.3%; risk difference, -21.9 percentage points [95% CI, -29.1 to -14.7]; p<0.001) (Figure 1A). The 5-year Kaplan-Meier survival estimates were 96.1% in the sotatercept group versus 61.2% in the control group (log-rank p=0.042), with a hazard ratio of 0.40 (95% CI, 0.16 to 0.99) (Figure 2A).

**Figure 1.**
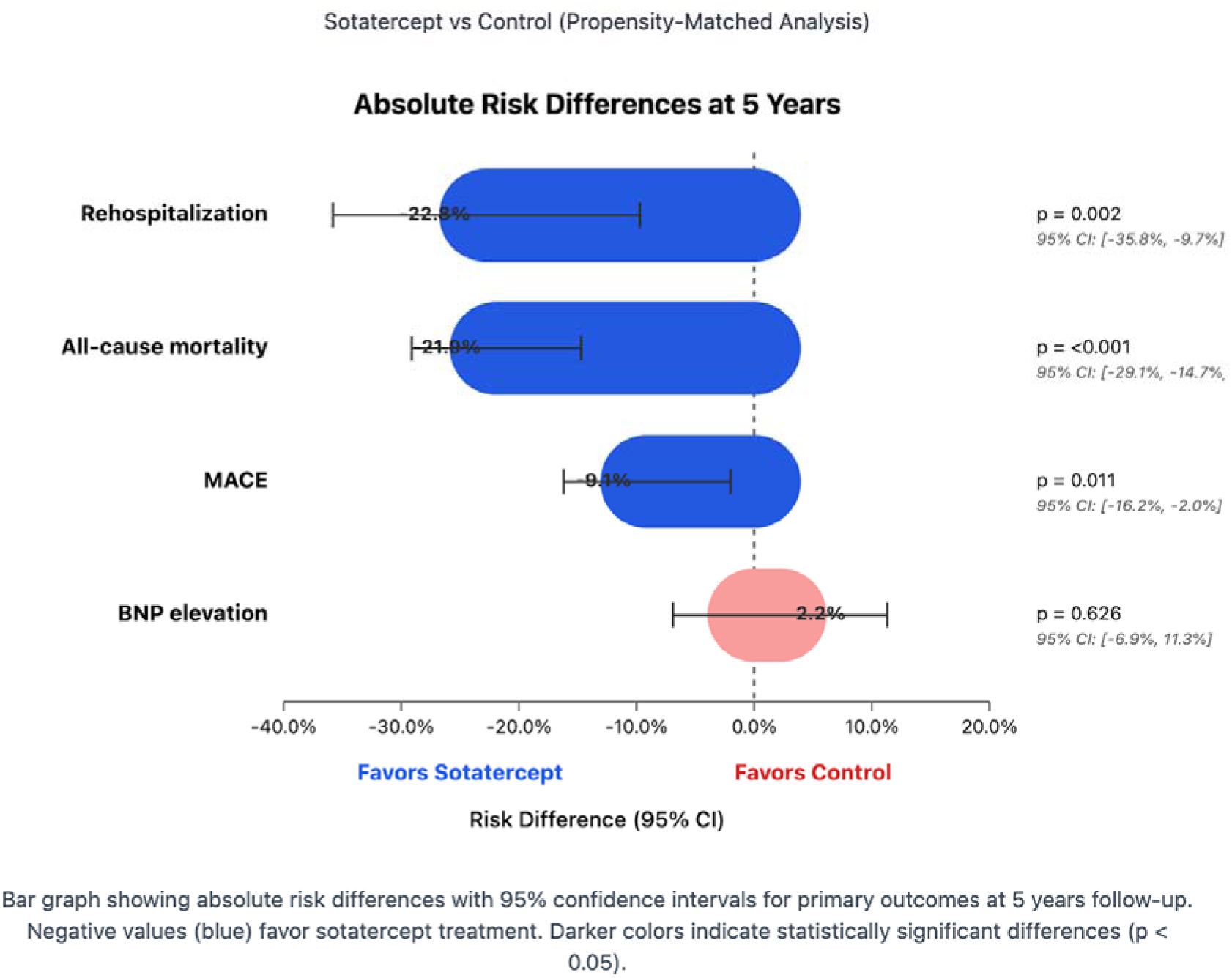
Risk Differences for Primary Outcomes at 5 Years.

**Figure 2.**
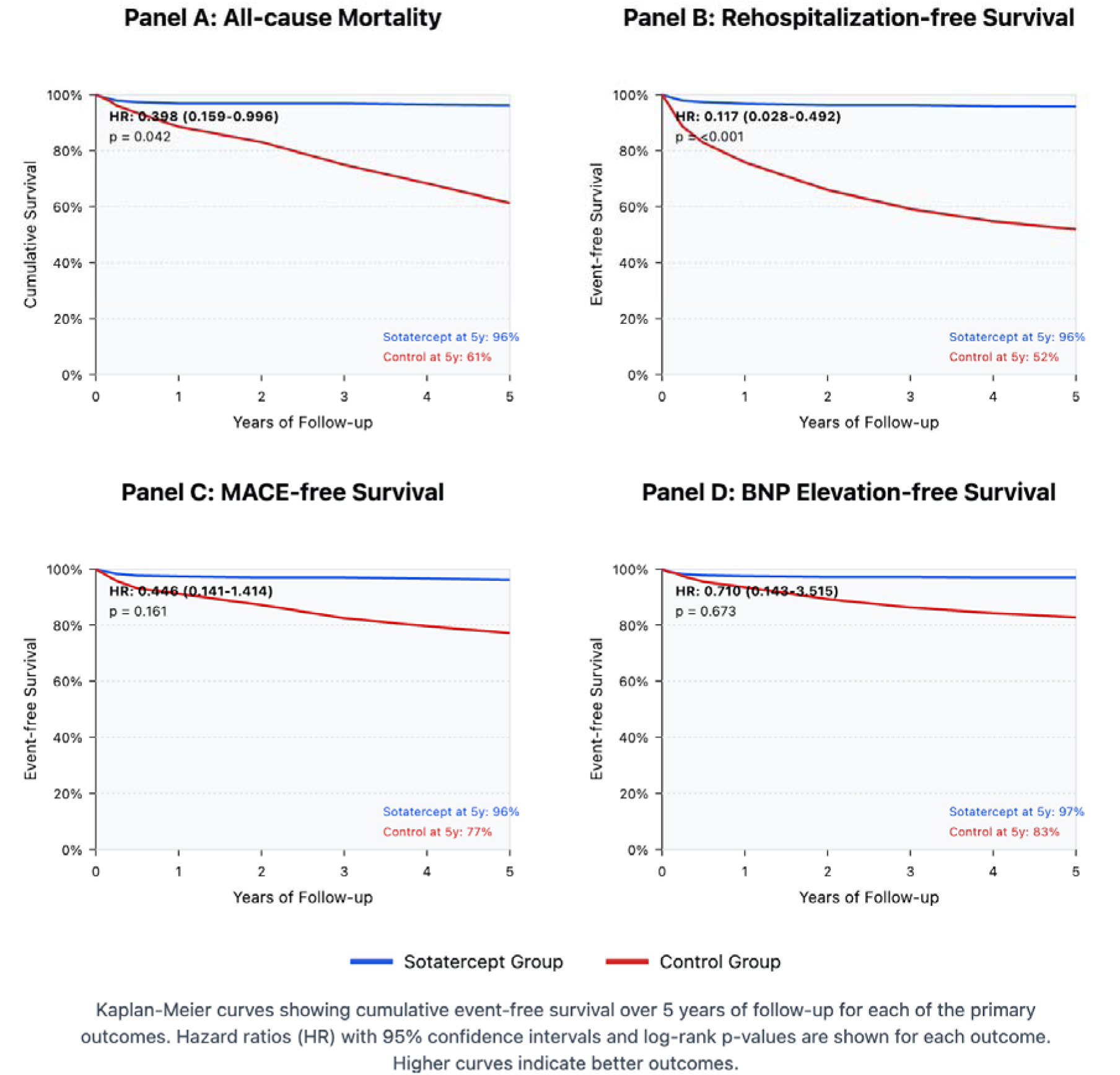
Kaplan-Meier Survival Curves (5-year follow-up) ⍰ Panel A: All-cause mortality ⍰ Panel B: Rehospitalization-free survival ⍰ Panel C: MACE-free survival ⍰ Panel D: BNP-event-free survival

At 3 years of follow-up, similar mortality benefits were observed (6.0% vs 21.6%; risk difference, -15.6 percentage points [95% CI, -22.8 to -8.4]; p<0.001), with Kaplan-Meier survival estimates of 97.4% versus 74.9% (log-rank p=0.015) and a hazard ratio of 0.28 (95% CI, 0.09 to 0.84). The mortality benefit was also observed, although with lower statistical significance, at 1-year follow-up (5.4% vs. 10.4%; risk difference, -5.0 percentage points [95% CI, -10.5 to 0.5]; p=0.076).

### Rehospitalization

Patients receiving sotatercept demonstrated significantly lower rehospitalization rates at 5 years compared with the control group (15.4% vs. 38.1%; risk difference, -22.8 percentage points [95% CI, -35.8 to -9.7]; p=0.002) (Figure 1B). Kaplan-Meier analysis confirmed a substantial reduction in hospitalization-free survival (log-rank p<0.001) with a hazard ratio of 0.12 (95% CI, 0.03 to 0.49) (Figure 2B). The mean number of rehospitalizations per patient was also lower in the sotatercept group compared with the control group (1.5 vs. 14.8 over 5 years).

At 3 years, rehospitalizations remained significantly lower in the sotatercept group (16.4% vs. 30.9%; risk difference, -14.5 percentage points [95% CI, -28.3 to -0.7]; p=0.025). The 1-year analysis demonstrated consistent findings (15.4% vs 24.7%; risk difference, -9.4 percentage points [95% CI, -21.6 to 2.9]; p=0.152).

### Major Adverse Cardiovascular Events

MACE occurrence at 5 years was significantly lower in the sotatercept group compared with controls (6.4% vs. 15.5%; risk difference, -9.1 percentage points [95% CI, -16.2 to -2.0]; p=0.011) (Figure 1C). Kaplan-Meier analysis showed improved MACE-free survival in the sotatercept group (log-rank p=0.161) with a hazard ratio of 0.45 (95% CI, 0.14 to 1.41). The mean number of MACE events per patient was 2.0 in the sotatercept group versus 3.0 in the control group over 5 years.

The 3-year analysis showed consistent MACE reduction (6.9% vs 11.0%; risk difference, -4.1 percentage points [95% CI, -10.8 to 2.6]; p=0.231), as did the 1-year analysis (6.4% vs 7.0%; risk difference, -0.6 percentage points [95% CI, -6.3 to 5.1]; p=0.828).

### BNP Levels

The proportion of patients with elevated BNP (≥100 pg/mL) during follow-up did not differ significantly between the sotatercept and control groups at 5 years (12.8% v.s 10.6%; risk difference, 2.2 percentage points [95% CI, -6.9 to 11.3]; p=0.626) (Figure 1D). Kaplan-Meier estimates of BNP-event-free survival were similar between groups (log-rank p=0.673) with a hazard ratio of 0.71 (95% CI, 0.14 to 3.52). This lack of significant difference in BNP events was consistent across all follow-up periods.

### Outcome Consistency Across Follow-up Periods

The benefit associated with sotatercept increased with longer follow-up for mortality, rehospitalization, and MACE outcomes (Figure 3). The most robust and consistent benefit was observed for rehospitalization, which maintained statistical significance across most time points. Risk ratios favoring sotatercept were consistently below 0.5 for all clinical outcomes except BNP at the 5-year follow-up.

**Figure 3.**
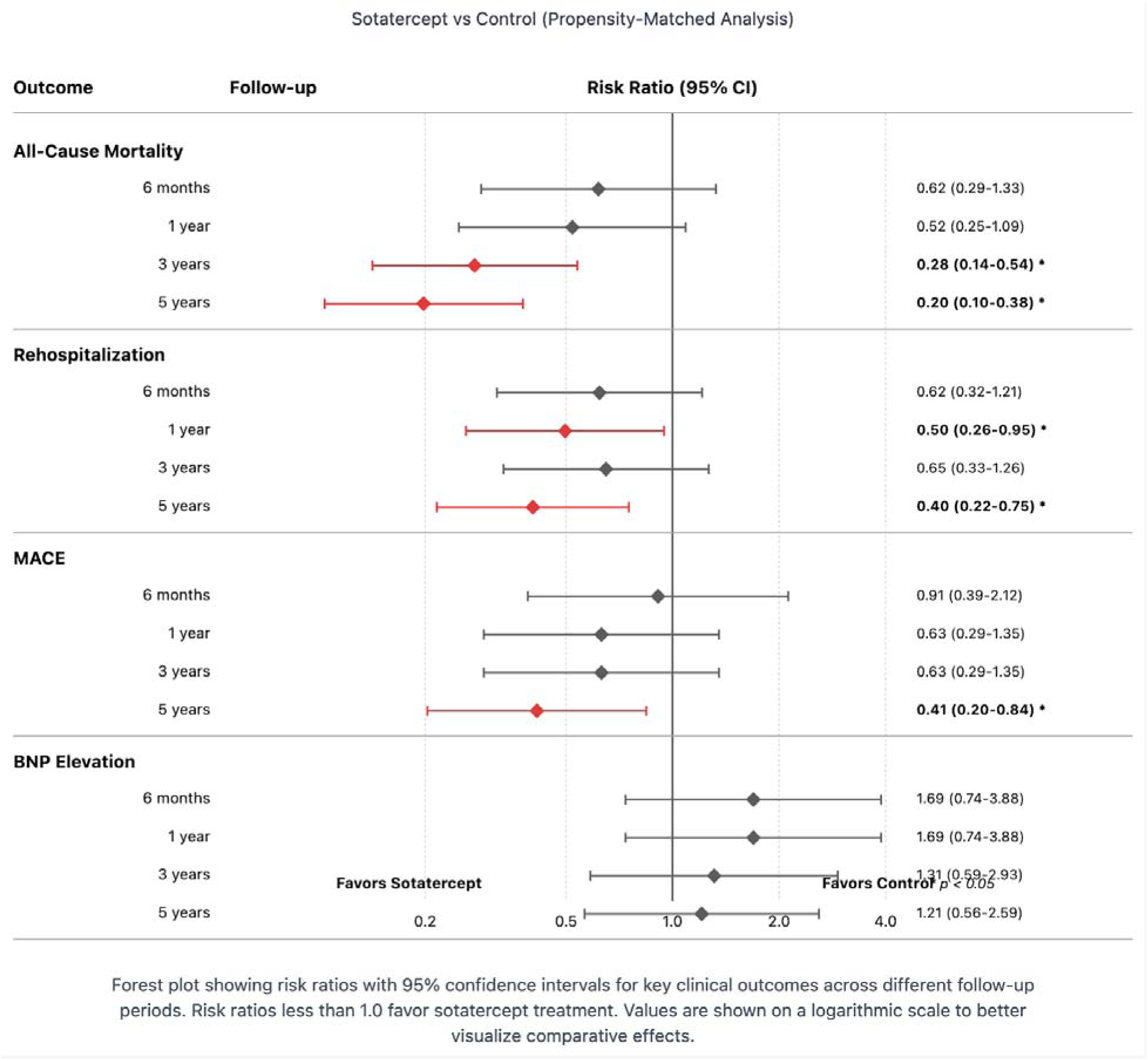
Risk Ratios Across Different Follow-up Periods. ⍰ Forest plot showing risk ratios with 95% CI for each outcome at: ⍰ 6 months ⍰ 1 year ⍰ 3 years ⍰ 5 years

## Discussion

Pulmonary hypertension (pHTN) is a progressive, life-threatening condition that is characterized by the excessive proliferation of pulmonary arterial tissue [21]. Treatments for this condition have primarily targeted the pathophysiology of the disease, including vasodilators and antiproliferative therapies aiming to slow the disease progression and manage quality of life [22]. Sotatercept is a novel, recently approved recombinant fusion protein that acts as an activin signaling inhibitor, reducing vascular proliferation by balancing pro-proliferative growth factor cascades [22,23]. Recent clinical trials, such as the PULSAR and STELLAR, have significantly improved pulmonary vascular resistance and exercise capacity at 24 weeks [5,6]. While these results are promising for short-term outcomes, very few studies have explored the long-term impact of this medication on clinical outcomes for patients with pHTN. Our retrospective cohort analysis examines the impact of sotatercept on clinical outcomes for patients with pHTN for up to 5 years, representing the extended follow-up on this therapy to date.

.hOur study also found that sotatercept significantly reduced rates of rehospitalization at 1 year and 5 years compared to the control. Rehospitalization is a significant issue for patients with pHTN, with the REVEAL registry reporting that among patients newly diagnosed with pHTN, 56.8% had at least one hospitalization post-enrollment, with over half being pHTN-related [28]. The authors found that pHTN hospitalization was associated with higher rehospitalizations and worse survival at 3 years [28].

Rehospitalization for pHTN patients has also been shown to have a higher length of stay and drive hospitalization costs, emphasizing its impact on the healthcare system in addition to the patients and their families [29]. An interesting finding in our study was that sotatercept did not significantly reduce the rehospitalization rate at 3 years, despite significant reductions at 1 year and 5 years. This could potentially highlight that the effects of sotatercept on rehospitalizations might be time-dependent, as shown in our study with mortality and MACE.

Finally, we found no significant difference between sotatercept and control in BNP elevation throughout our study period. This contrasts with the PULSAR and STELLAR trials, which showed a significant reduction in NT-proBNP at 24 weeks with sotatercept compared to placebo [5,6]. Some potential reasons for this disparity include our study exploring long-term outcomes of sotatercept and our control group being treated with pHTN therapies. In contrast, the PULSAR and STELLAR trials utilized a placebo [5,6]. However, the SOTERIA study, which also included a control group being treated with pHTN therapy, did report that reductions in NT-proBNP levels were maintained for one year [24]. While the stability in BNP levels shown in our study is promising, this disparity highlights the importance of future studies exploring the long-term impacts of sotatercept to improve our understanding of this therapy, especially as increased NT-proBNP levels have been associated with an increased risk of mortality and lung transplantation in patients with pHTN [30]. It is important to highlight that our study compared the impact of sotatercept on BNP levels in contrast to previous studies utilizing NT-proBNP levels. Most studies show no significant differences between either as a measure of pHTN. However, some suggest that BNP levels might correlate better with pulmonary hemodynamics, while NT-proBNP levels might correlate better with the prognosis for this disease [31].

### Limitations

It is important to acknowledge that our study carries several limitations. Our study’s retrospective, observational nature might limit its power and ability to conclude compared to randomized controlled trials. We also did not explore the long-term impact of sotatercept on functional parameters, such as a six-minute walking distance, which could have added helpful insights into its long-term efficacy. Prior to the FDA approval of sotatercept, between 2018 and 2024, its prescription in the United States was relatively limited. However, due to the nature of TrinetX, we were able to longitudinally track patients who were enrolled in clinical trials and received sotatercept during the investigational period. Additionally, we could not extract parameters such as pulmonary vascular resistance, WHO functional class, or quality of life measures, which the medication could have impacted. Despite these limitations, our study was able to explore the long-term clinical impacts of sotatercept to a follow-up period of 5 years, revealing promising benefits worth exploring in future clinical trials.

## Conclusions

Our study found that sotatercept may provide a progressive reduction in mortality, major adverse cardiovascular events, and rehospitalization for up to five years of follow-up, with no significant differences in BNP levels throughout our study period. This study represents the longest follow-up on sotatercept to date and highlights potential benefits worth exploring in future clinical trials.

## Data Availability

All data produced in the present work are contained in the manuscript

## Supplementary Appendix

**Supplementary Figure S1.**
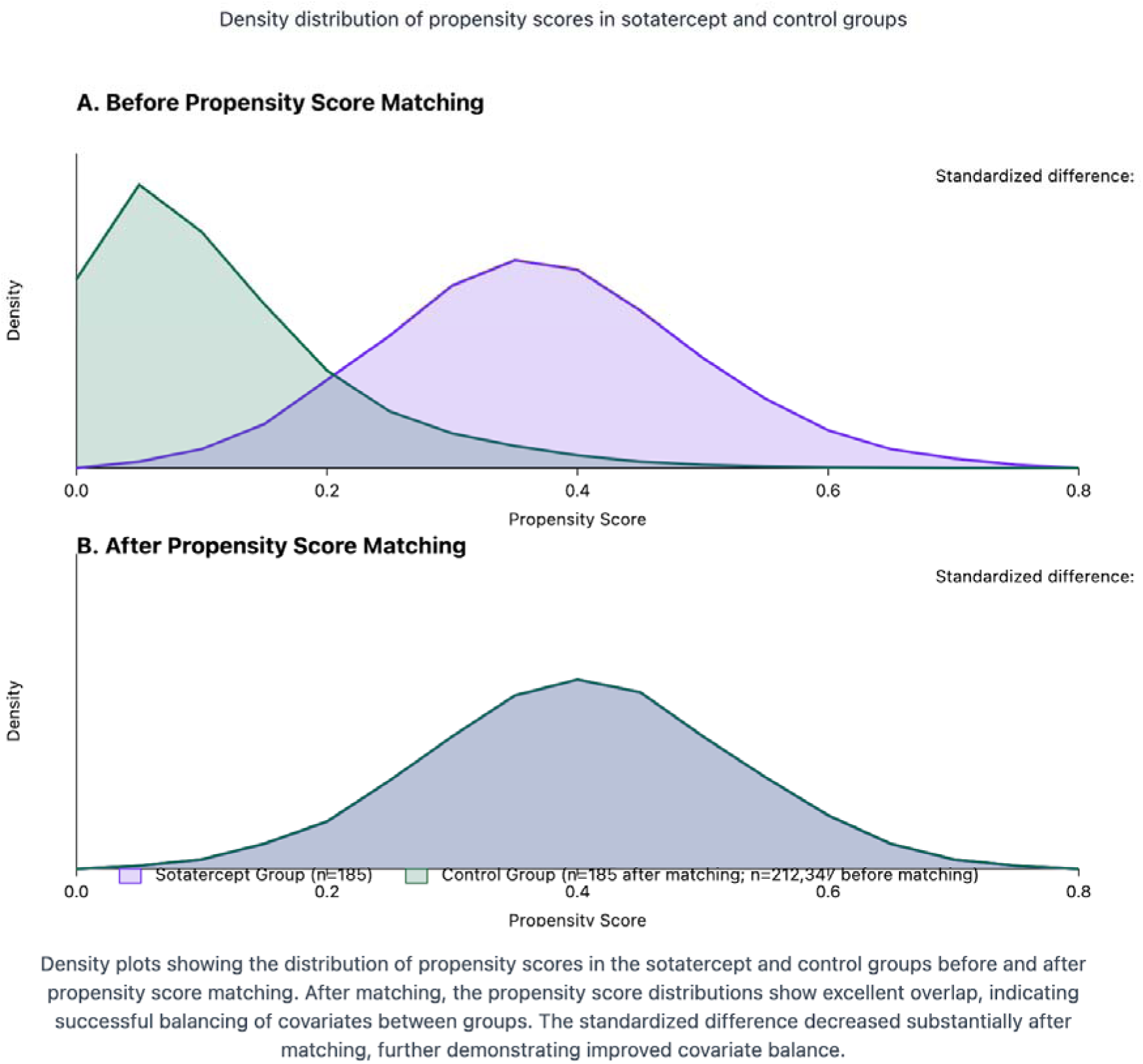
Propensity Score Distribution Before and After Matching. ⍰ Density plots showing improved overlap after matching

**Supplementary Table S1.**
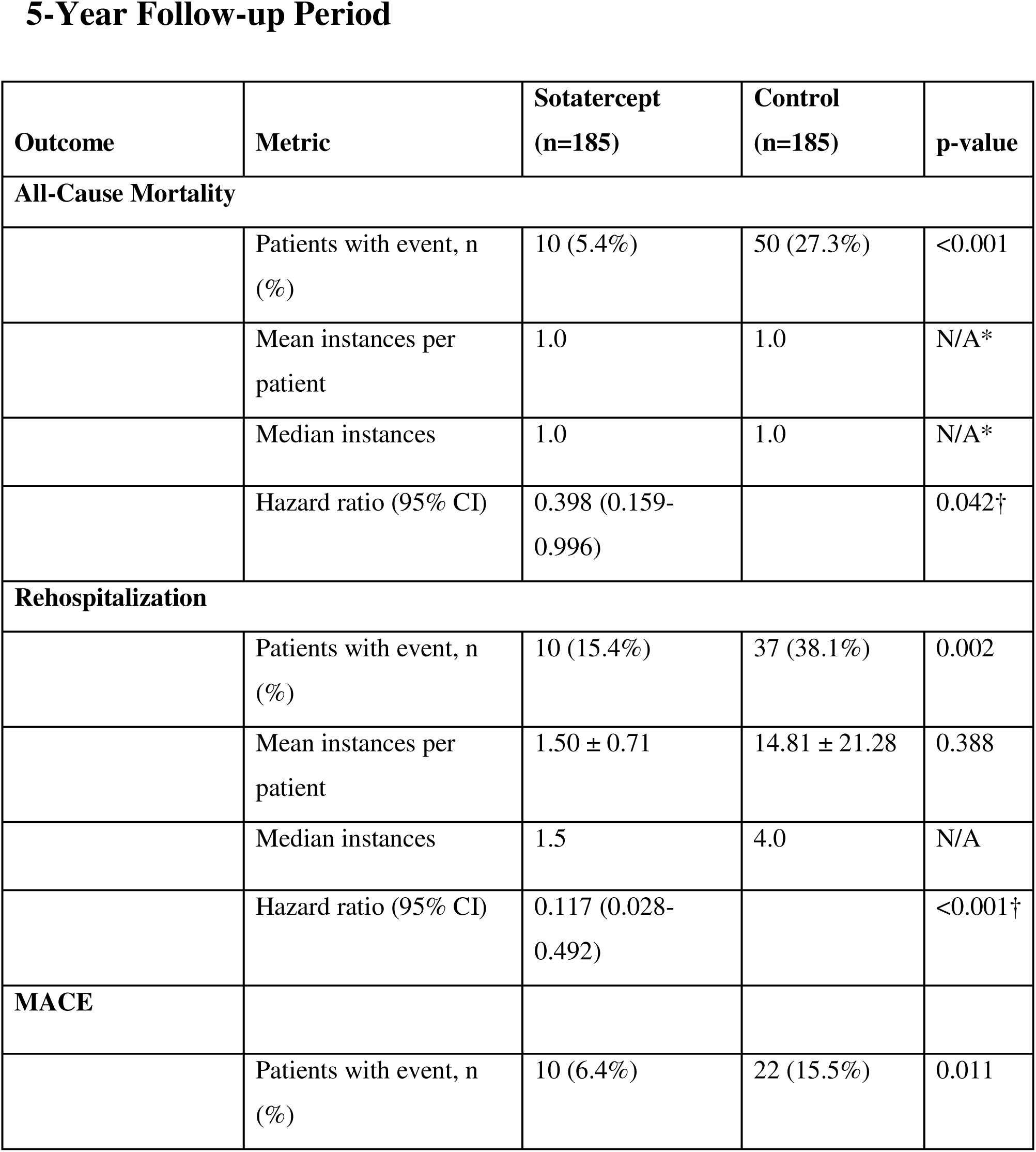
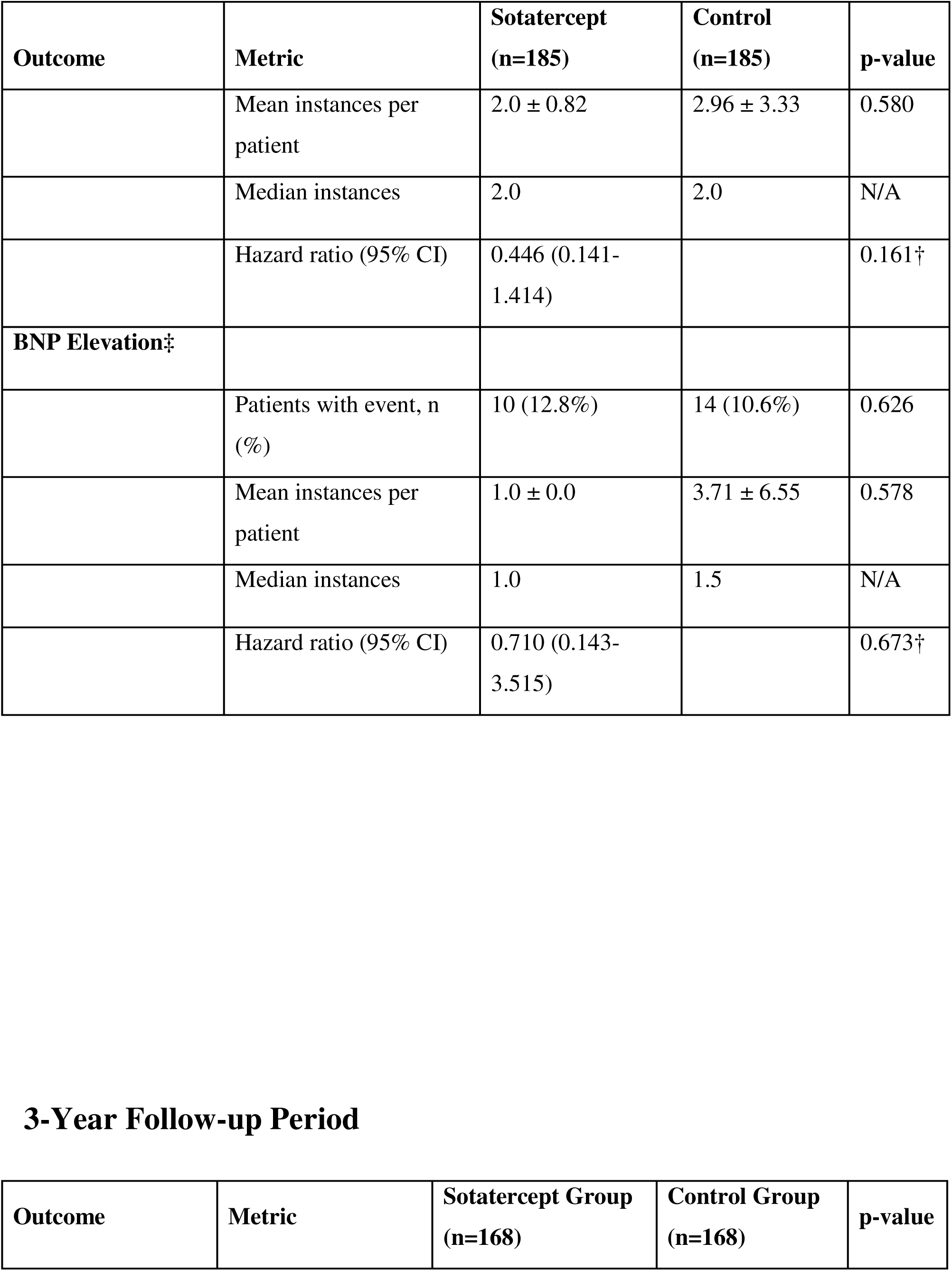
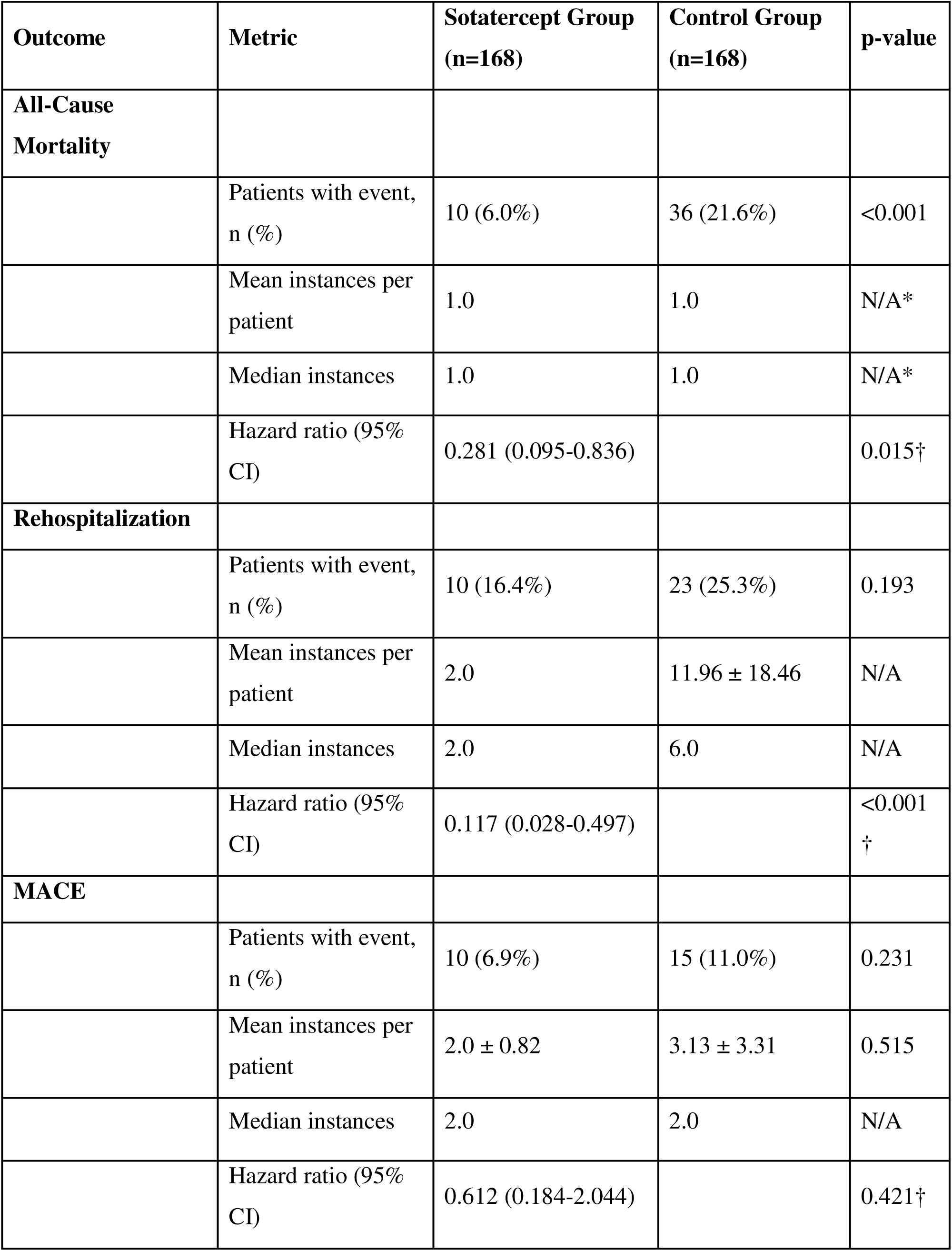
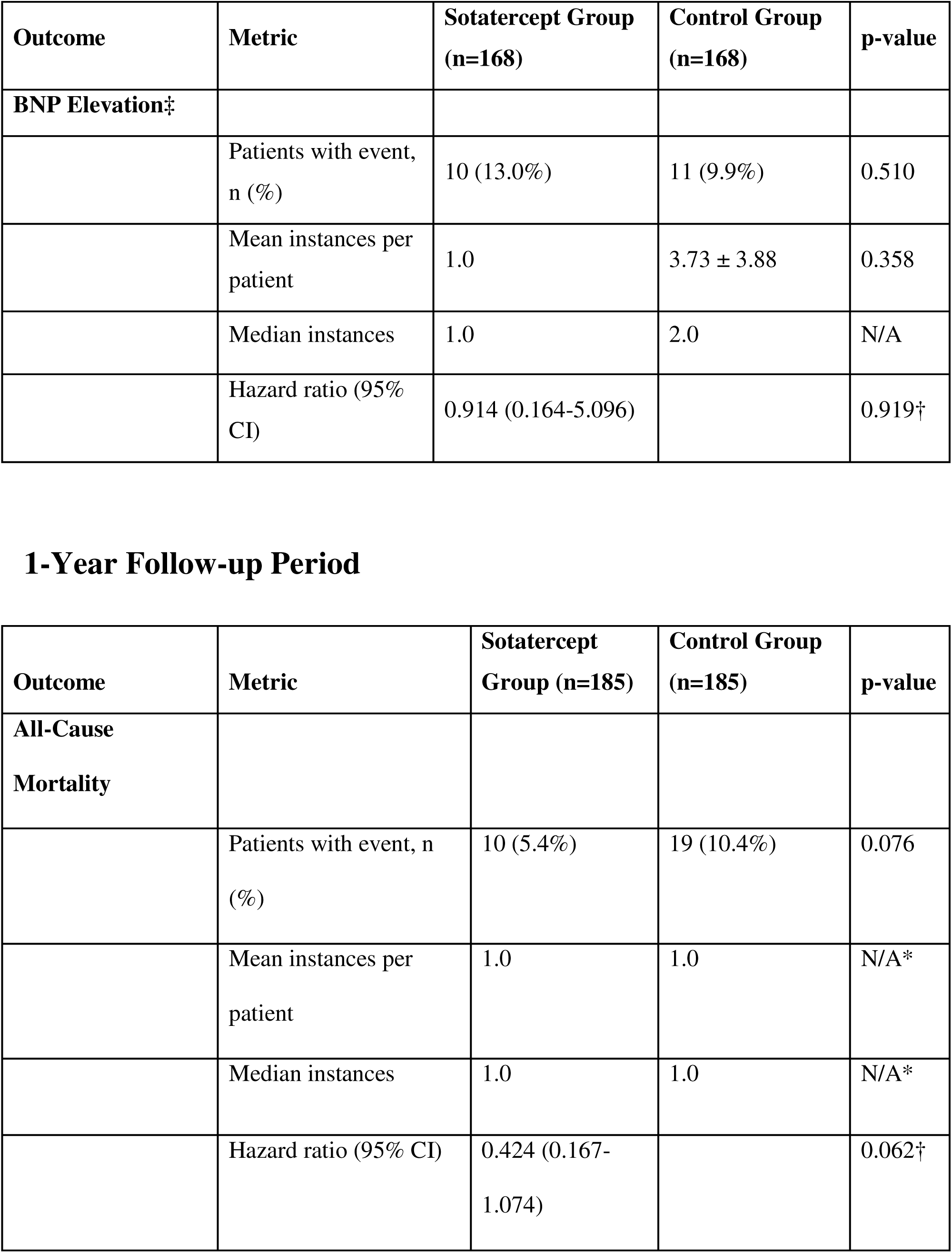
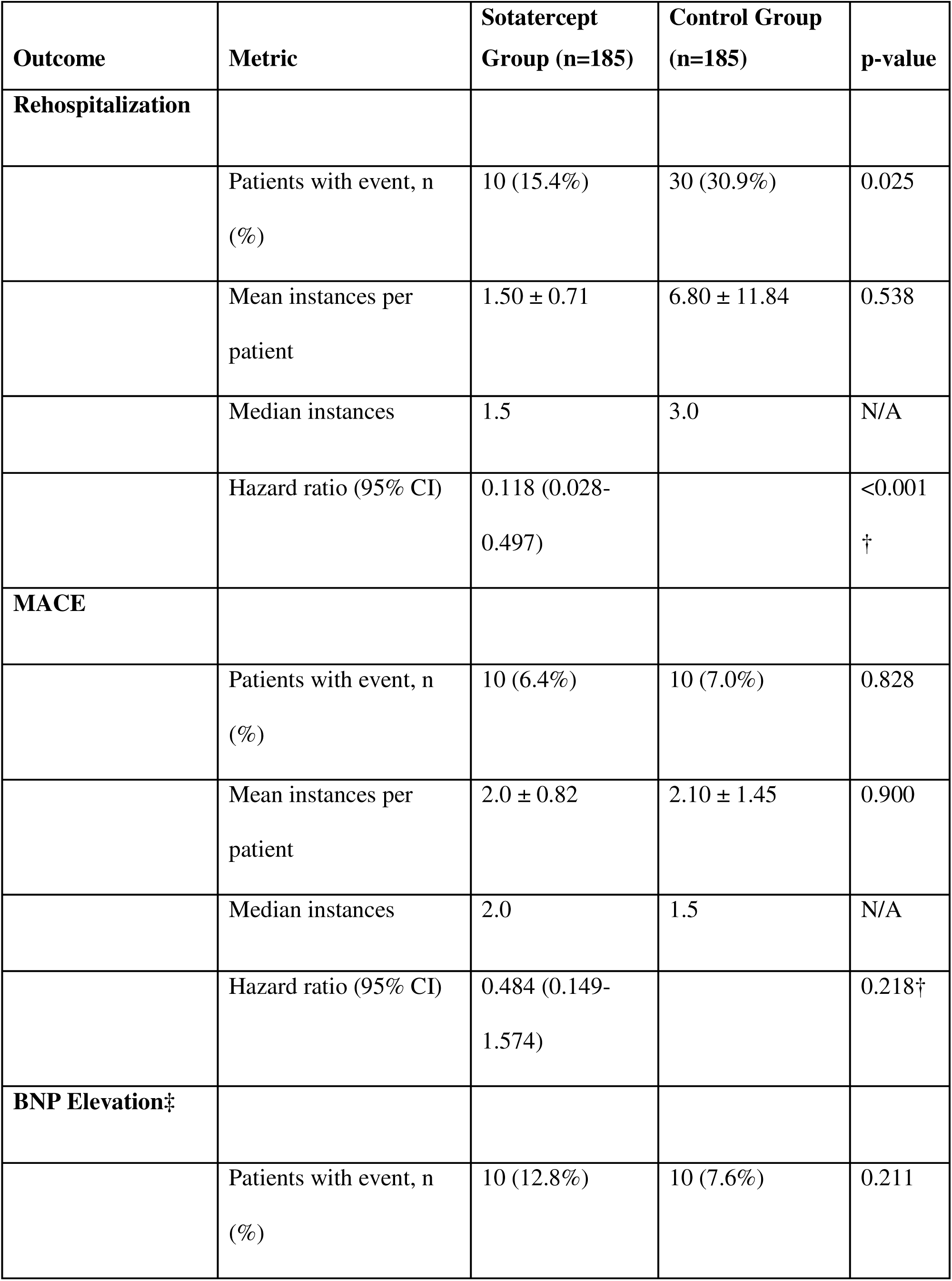
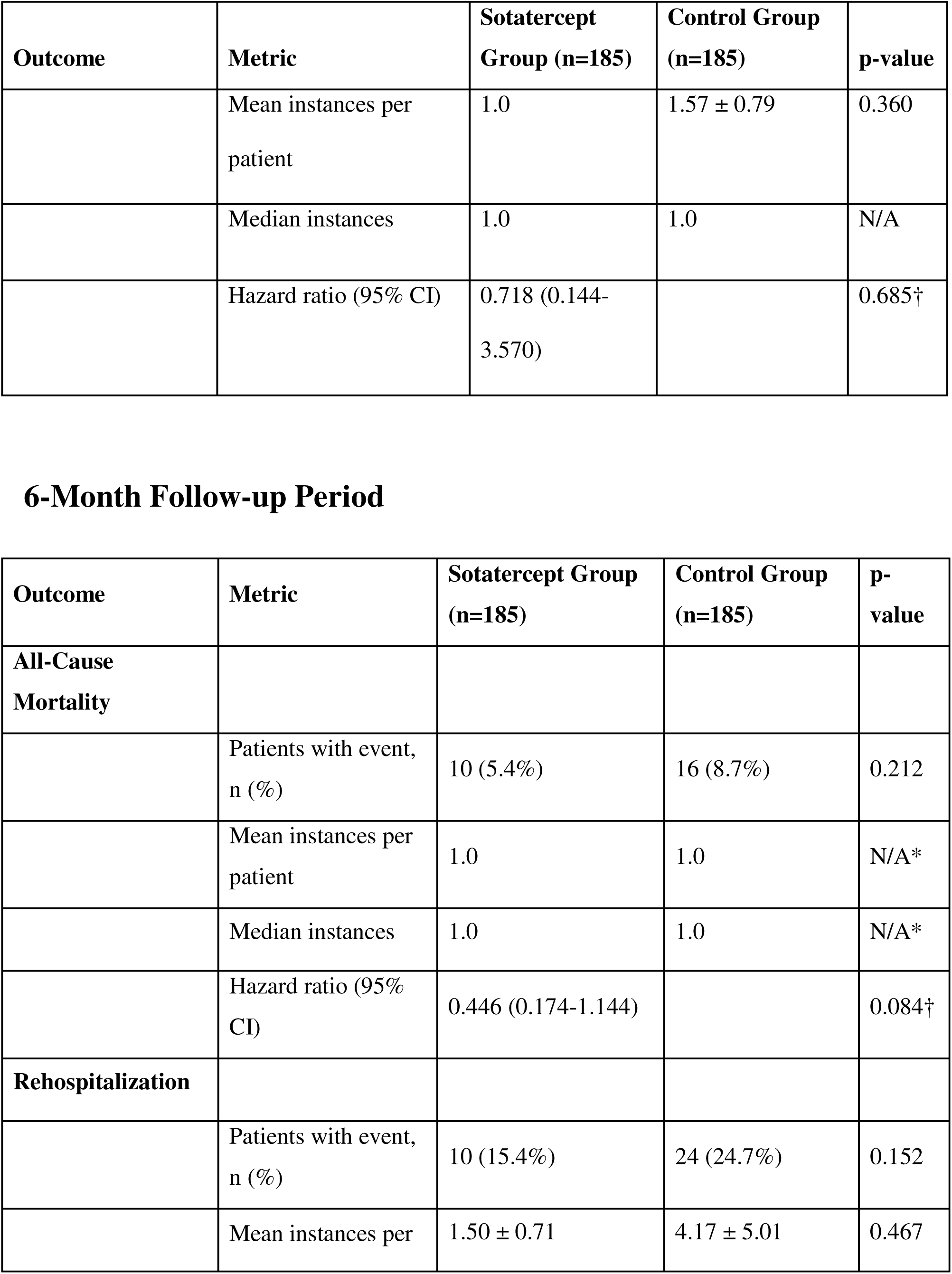
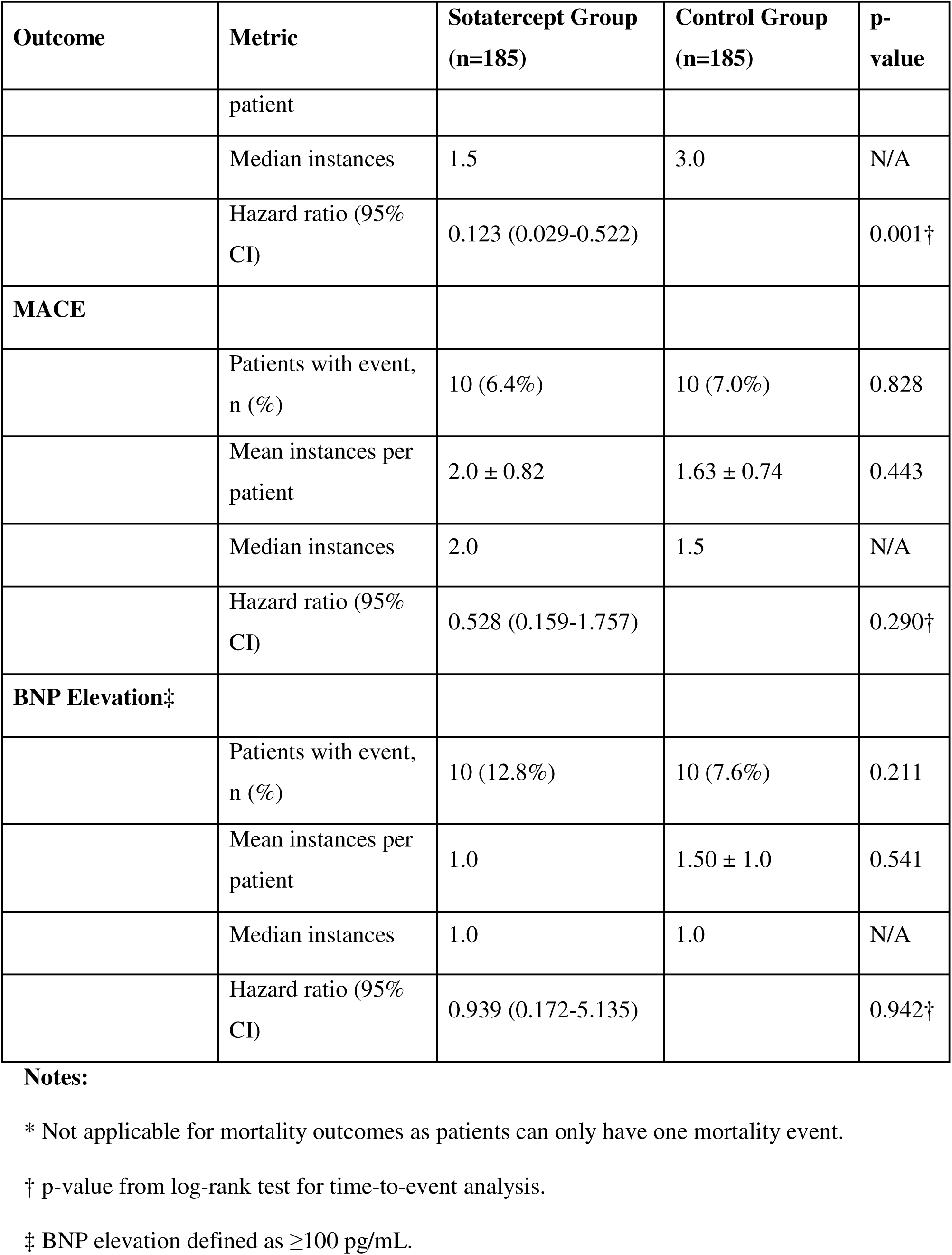
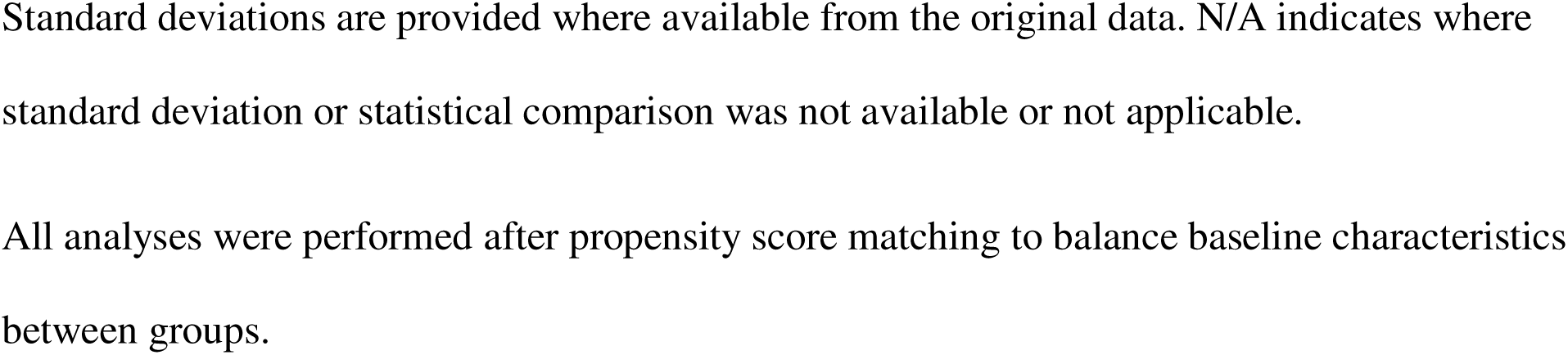
Number of Events and Instances by Outcome Category. ⍰ Detailed breakdown of event counts, mean numbers per patient ⍰ P-values for between-group comparisons

### Central Illustration. Comprehensive Summary of Sotatercept Benefits in Pulmonary Hypertension

⍰ Visual abstract integrating key findings across all outcomes and follow-up periods

⍰ Potential mechanistic framework for observed clinical benefits

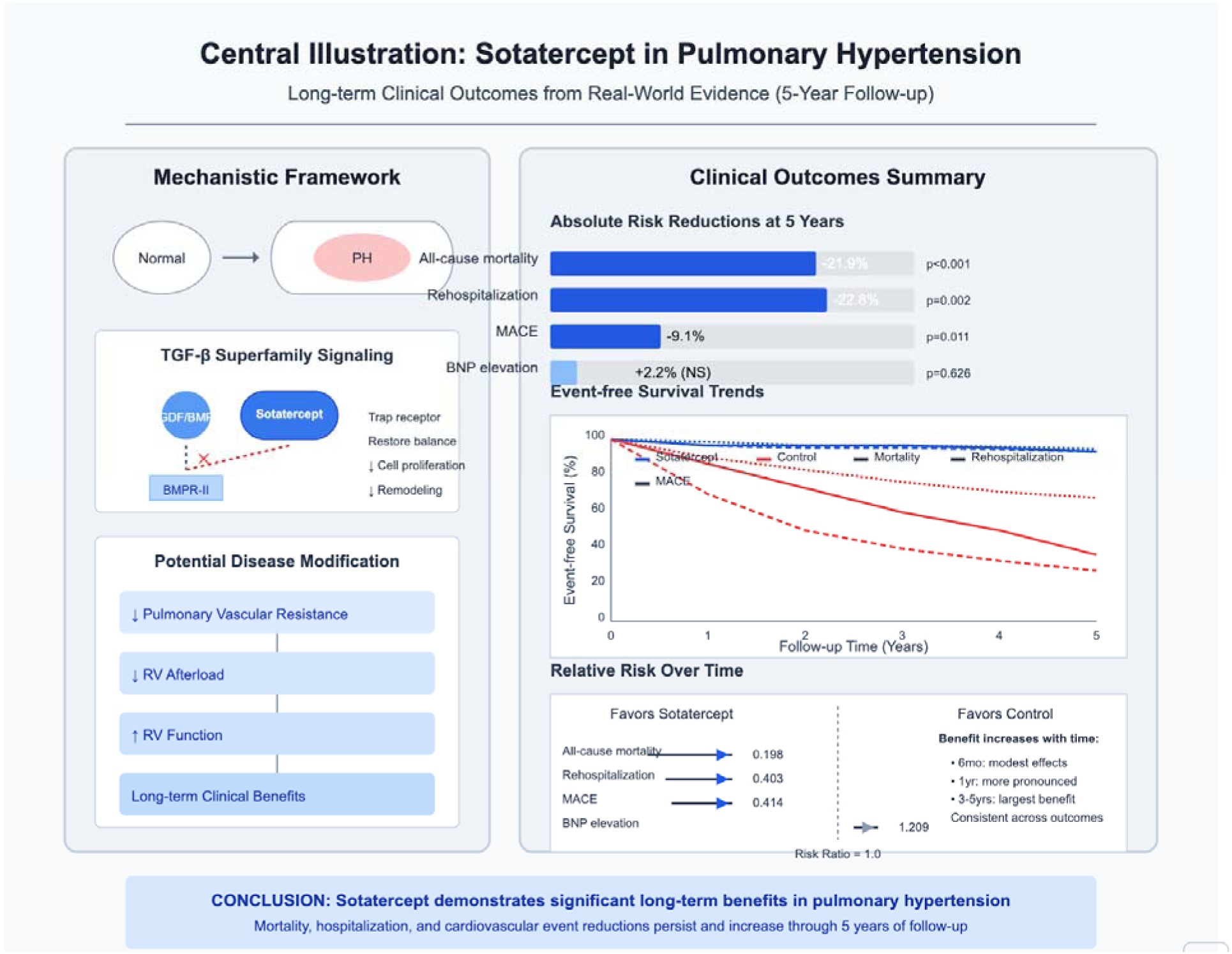

## References

1. Huang H, Hu C, Zhang R, Xu H, Cao M, Fu Y. Global Burden of Pulmonary Arterial Hypertension and Associated Heart Failure: Global Burden of Disease 2021 Analysis. JACC Heart Fail. 2025 Mar 5:102385. doi: 10.1016/j.jchf.2024.12.005.

2. Gelfman DM. Diagnosis: Pulmonary Hypertension. Next Steps. Am J Med. 2025 Jun;138(6):941-945. doi: 10.1016/j.amjmed.2025.01.028. Epub 2025 Feb 1. PMID: 39892488.

3. Liu L, Li C, Cai J, Kong R, Wang Y, Wang Y, Li S, Zhan J, Liu Y. Trends and levels of the global, regional, and national burden of pulmonary arterial hypertension from 1990 to 2021: findings from the global burden of disease study 2021. Front Med (Lausanne). 2024 Dec 10;11:1515961. doi: 10.3389/fmed.2024.1515961.

4. Farber HW, Miller DP, Poms AD, Badesch DB, Frost AE, Muros-Le Rouzic E, Romero AJ, Benton WW, Elliott CG, McGoon MD, Benza RL. Five-Year outcomes of patients enrolled in the REVEAL Registry. Chest. 2015 Oct;148(4):1043–54. doi: 10.1378/chest.15-0300. PMID: 26066077.

5. Hoeper MM, Badesch DB, Ghofrani HA, et al. Phase 3 Trial of Sotatercept for Treatment of Pulmonary Arterial Hypertension. N Engl J Med. 2023;388(16):1478–1490.

6. Humbert M, McLaughlin V, Gibbs JS, et al. Sotatercept for the treatment of pulmonary arterial hypertension. N Engl J Med. 2021;384(13):1204–1215.

7. Sahay S, Chakinala MM, Kim NH, et al. Contemporary Treatment of Pulmonary Arterial Hypertension: A U.S. Perspective. Am J Respir Crit Care Med. 2024;210(5):581–592.

8. Pitre T, Desai K, Mah J, et al. Comparative Effectiveness of Sotatercept and Approved Add-On Pulmonary Arterial Hypertension Therapies: A Systematic Review and Network Meta-Analysis. Ann Am Thorac Soc. 2024;21(8):1194–1203.

9. Stapff M, Hilderbrand S. Real[world evidence in cardiovascular medicine: assuring quality and relevance for scientific discovery and medical practice. J Am Heart Assoc. 2022;11(14):e026693.

10. Block JP, Chandra A, McManus DD, Willett WC. Implementing a real-world evidence ecosystem using federated data networks. N Engl J Med. 2023;388(1):12–17.

11. Khosla NG, Anand A, Lewnard JA. A comparison of patient characteristics and clinical outcomes in patients hospitalized with influenza before vs during the coronavirus disease 2019 (COVID-19) pandemic. JAMA Netw Open. 2023;6(10):e2338399.

12. Franklin JM, Patorno E, Desai RJ, et al. Emulating randomized clinical trials with nonrandomized real-world evidence studies: first results from the RCT DUPLICATE initiative. Circulation. 2021;143(10):1002–1013.

13. Rosenbaum PR, Rubin DB. The central role of the propensity score in observational studies for causal effects. Biometrika. 1983;70(1):41–55.

14. Austin PC. Optimal caliper widths for propensity-score matching when estimating differences in means and differences in proportions in observational studies. Pharm Stat. 2011;10(2):150–161.

15. Wang SV, Jin Y, Fireman B, et al. Relative performance of propensity score matching strategies for subgroup analyses. Am J Epidemiol. 2018;187(8):1799–1807.

16. Therneau TM, Grambsch PM. Modeling Survival Data: Extending the Cox Model. New York, NY: Springer; 2024.

17. Patel MR, Kabir M, Chari R, Zhang Z. Modern causal inference methods in real-world health databases: opportunities and applications for cardiopulmonary research. Chest. 2024;165(2):329–342.

18. Benjamini Y, Hochberg Y. Controlling the false discovery rate: a practical and powerful approach to multiple testing. J R Stat Soc Series B Stat Methodol. 1995;57(1):289–300.

19. Schneeweiss S, Rassen JA, Glynn RJ, et al. High-dimensional propensity score adjustment in studies of treatment effects using health care claims data. Epidemiology. 2009;20(4):512–522.

20. Baiocchi M, Cheng J, Small DS. Instrumental variable methods for causal inference. Stat Med. 2024;43(1):66–94.

21. Demerouti E, Frantzeskaki F, Adamidi T, et al. Revisiting treatment of pulmonary arterial hypertension in the current era: A Greek scientific document. Hellenic Journal of Cardiology. Published online February 2025.

22. Miranda AC, Cornelio CK, Tran BAC, Fernandez J. Sotatercept: A First-In-Class Activin Signaling Inhibitor for Pulmonary Arterial Hypertension. J Pharm Technol. Published online February 22, 2025.

23. Fujiwara T, Ishii S, Minatsuki S, et al. Exploring novel therapeutics for pulmonary arterial hypertension. International Heart Journal. 2025;66(1):3–12.

24. Preston IR, Badesch D, Ghofrani H-A, et al. A long-term follow-up study of Sotatercept for treatment of pulmonary arterial hypertension: Interim results of soteria. Eur Respir J. Published online February 20, 2025:2401435.

25. Hoeper MM, Pausch C, Grünig E, et al. Temporal trends in pulmonary arterial hypertension: results from the COMPERA registry. Eur Respir J. 2022;59:2102024.

26. Smilowitz NR, Armanious A, Bangalore S, et al. Cardiovascular Outcomes of Patients With Pulmonary Hypertension Undergoing Noncardiac Surgery. Am J Cardiol. 2019;123(9):1532–1537.

27. Zou T, Chen Q, Zhang L, et al. Pulmonary artery pressure is associated with mid-term major adverse cardiovascular events and postprocedure pericardial effusion in atrial fibrillation patients undergoing left atrial appendage occlusion. Ann Transl Med. 2021;9(16):1324.

28. Burger CD, Long PK, Shah MR, et al. Characterization of first-time hospitalizations in patients with newly diagnosed pulmonary arterial hypertension in the reveal registry. Chest. 2014;146(5):1263–1273.

29. Canavan N. Rehospitalization is driving costs in pulmonary arterial hypertension. Am Health Drug Benefits. 2013;6(9):600–601.

30. Hendriks PM, van de Groep LD, Veen KM, et al. Prognostic value of brain natriuretic peptides in patients with pulmonary arterial hypertension: A systematic review and meta-analysis. American Heart Journal. 2022;250:34–44.

31. Lewis RA, Durrington C, Condliffe R, Kiely DG. BNP/NT-proBNP in pulmonary arterial hypertension: time for point-of-care testing? Eur Respir Rev. 2020;29(156):200009.

